# Robust deep learning estimation of cortical bone porosity from MR T1-weighted images for individualized transcranial focused ultrasound planning

**DOI:** 10.1101/2024.07.18.24310644

**Authors:** Matthieu Dagommer, Mohammad Daneshzand, Aapo Nummemnaa, Bastien Guerin

**Affiliations:** École Supérieure de Physique et de Chimie Industrielles de la Ville de Paris (ESPCI), Paris France; Harvard Medical School, Boston MA; Athinoula A. Martinos Center for Biomedical Imaging, Massachusetts General Hospital, Charlestown MA

**Keywords:** <u>Keywords</u>: Deep learning, neural network, MRI, image-to-image translation, porosity, transcranial focused ultrasound

## Abstract

**Objective:** Transcranial focused ultrasound (tFUS) is an emerging neuromodulation approach that has been demonstrated in animals but is difficult to translate to humans because of acoustic attenuation and scattering in the skull. Optimal dose delivery requires subject-specific skull porosity estimates which has traditionally been done using CT. We propose a deep learning (DL) estimation of skull porosity from T1-weighted MRI images which removes the need for radiation-inducing CT scans.

**Approach:** We evaluate the impact of different DL approaches, including network architecture, input size and dimensionality, multichannel inputs, data augmentation, and loss functions. We also propose back-propagation in the mask (BIM), a method whereby only voxels inside the skull mask contribute to training. We evaluate the robustness of the best model to input image noise and MRI acquisition parameters and propagate porosity estimation errors in thousands of beam propagation scenarios.

**Main results:** Our best performing model is a cGAN with a ResNet-9 generator with 3D 64×64×64 inputs trained with L1 and L2 losses. The model achieved a mean absolute error of 6.9% in the test set, compared to 9.5% with the pseudo-CT of Izquierdo et al. (38% improvement) and 9.4% with the generic pixel-to-pixel image translation cGAN pix2pix (36% improvement). Acoustic dose distributions in the thalamus were more accurate with our approach than with the pseudo-CT approach of both Burgos et al. and Izquierdo et al, resulting in near-optimal treatment planning and dose estimation at all frequencies compared to CT (reference).

**Significance:** Our DL approach porosity estimates with ∼7% error, is robust to input image noise and MRI acquisition parameters (sequence, coils, field strength) and yields near-optimal treatment planning and dose estimates for both central (thalamus) and lateral brain targets (amygdala) in the 200-1000 kHz frequency range.

## 1. Introduction

Transcranial focused ultrasound (tFUS) is an emerging neuromodulation technique that may offer advantages compared to transcranial magnetic stimulation (TMS) as it can be focused deep in the brain with millimeter precision [1–3]. TFUS neuromodulation has been demonstrated in animals [4–9], but translation to humans is slow due, in part, to absorption and scattering in the thick human skulls that distort the beam and introduce deviations from the ideal, straight trajectory [10–12]. Although this problem is recognized as a major barrier to tFUS dose delivery, most ongoing tFUS neuromodulation clinical trials and studies ignore it and instead use line-of-sight (LOS) targeting, which assumes acoustic propagation along a straight line, thus likely resulting in sub-optimal dose delivery. This is a significant problem and raises the concern that ongoing human studies using LOS may not be conclusive. Finite-difference time domain [13–16], hybrid angle spectrum [12,17–23] or pseudo-spectral modeling [24–26] have been used for modeling of acoustic propagation through bone, however all those tools require individualized maps of the speed-of-sound and acoustic attenuation parameters as inputs. Those have traditionally been estimated using CT [27–29], but this can be difficult to justify as this increases radiation exposure and introduces cross-modality registration errors when using MRI for neuronavigation.

Estimation of acoustic maps from MRI can be done using zero echo-time (ZTE) MRI which mimics CT contrast and works well but requires specialized sequences [30–33]. A simpler approach is to generate a so-called pseudo-CT (pCT) map by non-linear registration of a population-average CT atlas into the subject’ frame, which only requires a standard clinical T1-weighted (T1w) MR image [34,35]. The main issue with this population-average approach is that it ignores large variations of bone porosity and density observed within and across individuals, see for example Fig. 1 and Ref. [11]. A more individualized approach consists in using deep learning (DL) of CT intensities from MR images. Liu et al. [36] proposed a 3D cGAN UNet for MRI-to-CT translation which they evaluated by reporting skull density ratio (SDR) as well as acoustic beams examples computed with Kranion [37] and k-wave [25]. Miscouridou et al. [38] proposed a similar model, however trained on ZTE MRI images, which they found was superior to T1w-based training for the task of tFUS beams estimation via k-wave. This echoes previous results by Su et al. [39], who deployed a 2D UNet for the estimation of CT Housfield units from ZTE MR images which they evaluated in estimations of SDR and beam profiles. More recently, Yaakub et al [40] and Koh et al [41] proposed similar 3D cGAN UNet models for estimation of Hounsfield units from T1w MR images, which they successfully evaluated in a limited number of k-wave simulations.

**Fig. 1.**
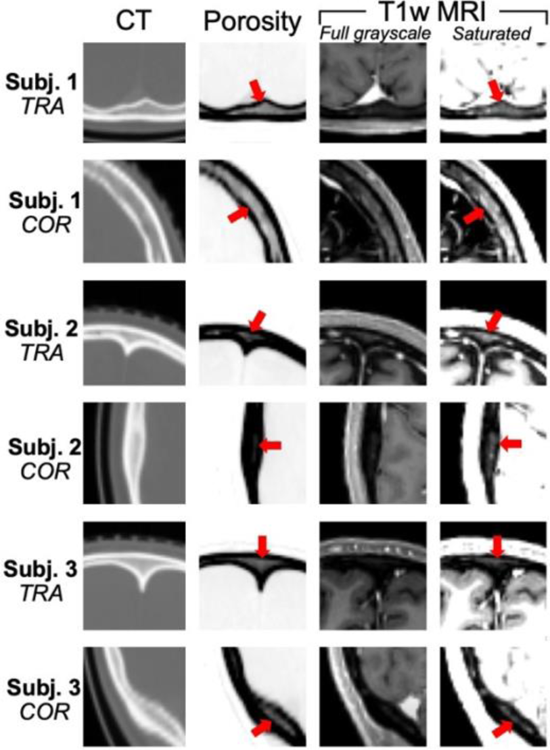
CT, porosity and T1-weighted MRI image sections for 3 of the 15 subjects included in this study. The porosity map is derived from CT as explained in the text. Red arrows point to high-porosity features corresponding to voxels containing primarily bone marrow, which are visible on both the porosity map and the saturated T1-weighted MR image. In other words, the T1-weighted MR image contain the information necessary to extract the porosity information in a patient-specific manner but doing so is not straightforward because of complex MRI contrast around the grey matter, white matter and the dura.

In this work, we propose a DL strategy for subject-specific estimation of the skull porosity from T1w MR images for the specific goal of improving the placement of focused transducers in tFUS neuromodulation studies. Our network is similar to that of Koh et al. and Yaakub et al., however with small but important differences. First, we optimize most aspects of the network (network architecture, input size/dimensionality, multichannel inputs, data augmentation, loss functions) in order to optimize performance. Second, we introduce backpropagation in the mask (BIM), a backpropagation approach that only considers pixels located inside the skull mask during training in an effort to focus the DL degrees-of-freedom on those pixels of maximum importance. In addition to improving performance, we show that limiting the estimation process to the skull mask makes the estimation robust to input image noise and variations in the various MRI acquisition parameters (sequence, coils). We compute the skull mask using SAMSEG [42], which is itself a robust method. As a results, the final model works even in the presence of very high noise in the input image and using T1-weighted MR images acquired at different field strength and using different sequences parameters and receive coils than for the training and validation sets. We evaluate our approach in simulations of thousands of transducer positions placed around the scalp of the test subject at 250, 500 and 1000 kHz (these are common frequencies used for neuromodulation). We distribute our code and trained weights online in an effort to disseminate our approach and improve reproducibility (https://github.com/bastpg).

## 2. Materials and methods

### 2.1 Network

Fig. 2 shows various DL configurations tested in this work. We used a conditional generative adversarial network (cGAN) architecture with UNet and ResNet generators and a patchGAN discriminator [43] (Fig. S1). We trained three 2D UNets for input sizes 256×256 (7 downsampling blocks), 128×128 (6 downsampling blocks) and 64×64 (5 downsampling blocks), as well as a 3D UNet for 64×64×64 inputs (Fig. 2B). We also trained a ResNet with 9 residual blocks (ResNet-9) both for 2D (256×256) and 3D inputs (64×64×64). Porosity outputs have the same size and dimensionality as the T1w image inputs, and the patchGAN discriminator was adapted to the size and dimensionality of those inputs/outputs. In addition to the MRI T1w input, the 256×256 2D Unet was also trained with an additional pCT channel (Izquierdo et al.) to assess whether this additional information improves convergence and prediction accuracy (Fig. 2C). We also implemented auto-context modeling whereby successive cGANs were daisy-chained together by using the output of the previous model as a channel input to the next, since this has been shown to be beneficial in some applications (Fig. 2D) [44,45].

**Fig. 2.**
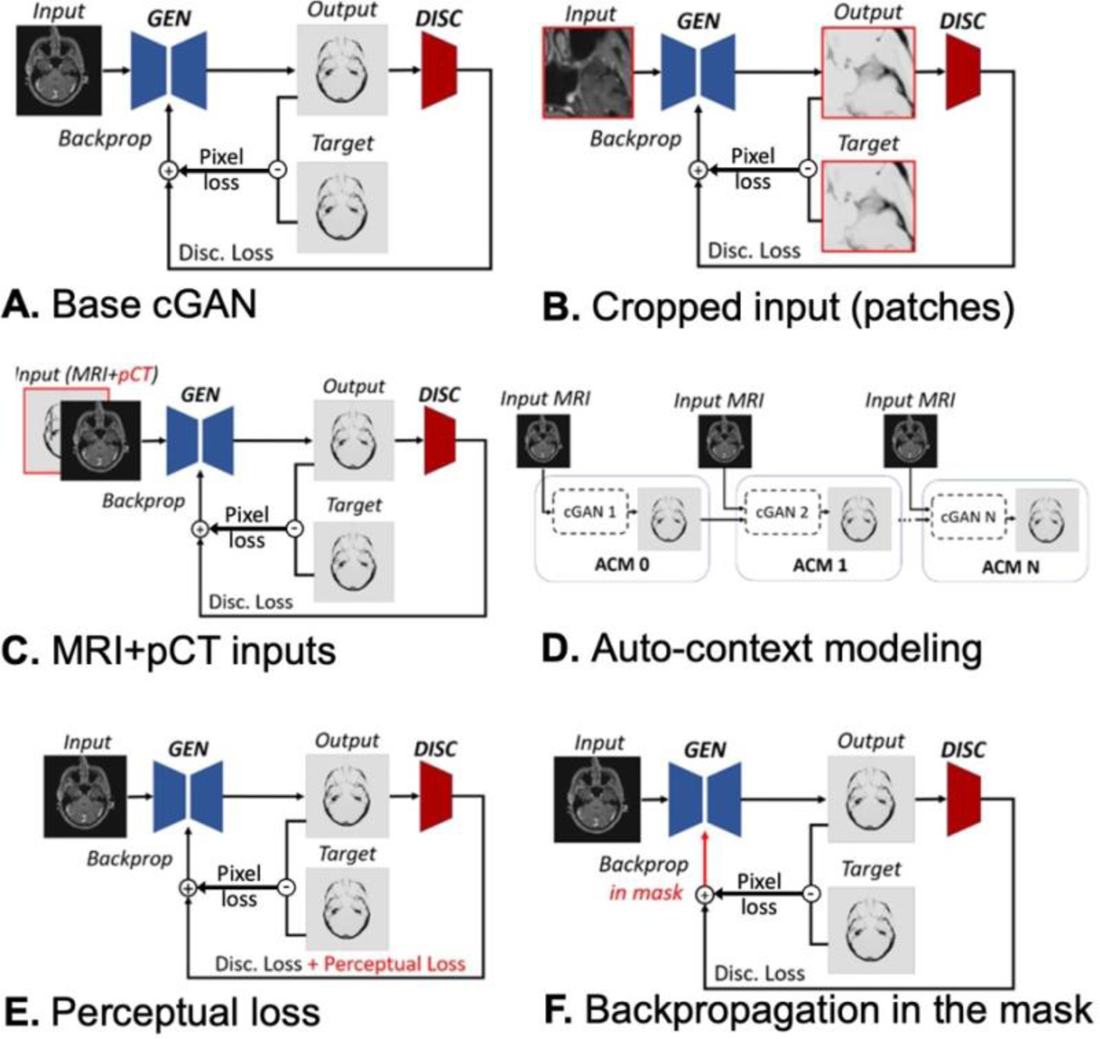
Deep learning configurations tested in this work. **A:** The base DL model used throughout this work is a conditional generative adversarial network (cGAN) with pixel and discriminator losses. All other models are based on this cGAN, with variations emphasized in red. **B:** cGAN with cropped inputs (size tested: 64×64, 128×128 and 256×256 for 2D, 64×64×64 for 3D). Cropping inputs yields a smaller generative network and increases the size of the training dataset, thus reducing overfitting. **C:** Use of the pseudo-CT as an additional input channel. **D:** Auto-context models are obtained by daisy-chaining cGANs together, which may improve the accuracy of the estimation at the last stage. **E:** Addition of perceptual loss to the backpropagation, in addition to the cGAN pixel and discriminator losses. **F:** Backpropagation in the mask only includes pixels located inside the skull mask in the backpropagation process. This is done in an effort to focus the degrees-of-freedom of the network on estimation of skull voxels, since porosity outside the mask is known and trivial (=1).

Next, we quantified the impact of the following loss functions: L1, L2, L1+L2, gradient difference loss (GDL) and perceptual loss. GDL penalizes difference between the x, y and z gradients of the estimated output and training reference, which has been shown to penalize edge discrepancies and tends to yield sharper image results [46]. Perceptual loss (Fig. 2E) was computed from intermediary features of the discriminator down-sampling layers thus yielding a richer, more complete picture of the differences between estimation output and reference [47]. Finally, we assessed backpropagation in the mask (BIM, Fig. 2F), whereby only pixels located inside the skull mask are included in the backpropagation process in an effort to focus the network’ degrees-of-freedom on the most important pixels, since pixels outside the mask have a trivial porosity value of one. The skull mask itself was estimated from the T1w MRI input using SAMSEG/Freesurfer [42]. For discriminator loss, we use an element-wise binary-cross entropy error term applied to the output of patchGAN.

### 2.2 Training, validation and testing

We used pairs of MRI and CT scans acquired in 15 different subjects for training, validation and testing [34]. We followed the basic rule of data segregation and used separate subjects for training (13 subjects), validation (1 subject) and testing (1 subject). Note that a single subject yields many inputs and outputs in the form of multiple slices, orientations (transverse, coronal, sagittal) and 3D patches. T1w MRI volumes were 1 mm isotropic MPRAGE acquired on a 3T Siemens ‘Skyra’ scanner (Siemens Healthineers, Erlangen Germany) minimally processed with intensity shading removal and interpolation in a standard 256×256×256 coordinate frame with 1 mm resolution [48]. The CT volumes were clinical scans acquired on a LightSpeed system (GE Healthcare, Chicago USA) with 0.45 mm resolution in-plane (transverse) and 2.5 mm slice thickness. Those were aligned and interpolated on the same 256×256×256 1 mm isotropic MRI grid using rigid registration [49]. Pseudo-CTs (pCT) were also generated for each subject following Ref. [34,35]. CT and pCT Hounsfield units (HU) were then converted into porosity maps *p* using the formula *p* = 1 − *HU*/1000 followed by normalization to [0;1] [50]. Slices below the nose were not included in training, validation and testing, since these locations are not relevant for tFUS. All inputs were normalized to [-1;1], thus matching the range of the tanh activation function used in the first layer of the networks used in this work.

Models were trained using a Tesla P100 PCIe GPU card with 16 GB of RAM using Adam optimization with learning rate = 0.0002, β_1_ = 0.5, β_2_ = 0.999 and ∈ = 10^−7^. All dropout layers were assigned a rate of 0.5 and the number of samples per epoch was set to 6149. Drawing with discount 473 random slices was used for both the validation and test subjects.

### 2.3 Volume reconstruction from patch outputs

When using cropped inputs, whole field-of-view outputs were reconstructed by tiling individual cropped outputs. 3D models of size 64×64×64 were obtained by cropping the original 256×256×256 input with a stride of 32 in all three directions. Reconstructions were obtained by averaging the cropped volumes in their intersections. For the 2D 256×256 models, transverse, coronal, and sagittal orientations were estimated independently and then averaged together.

### 2.4 Robustness to noise and MRI acquisition parameters

We evaluated the robustness of our approach to noise by adding increasing levels of Gaussian noise to the real and imaginary parts of the input MR images, thus resulting in the typical Rician noise distribution of MR magnitude data. The proposed DL estimation was then run on those datasets, including the SAMSEG segmentation step used to extract the skull mask. We also ran the proposed approach on six subjects from the 3 Tesla Human Connectome Project (young adult database, MGH sub-folder) [51] and 1.5 Tesla CERMEP [52] open-source repositories. Details of the MRI acquisitions (sequences, coils), which can be found in the Connectome and CERMEP references, were different from those used for the training, validation and test data. Importantly, the HCP data was acquired at 3T while the CERMEP data at 1.5T (training, validation and test images were acquired at 3T).

### 2.5 Acoustic simulations

The scalp surface of the test subject was meshed using a combination of MATLAB and iso2mesh [53] routines, resulting in 4868 faces after removal of mesh elements located below the ear and nose. A 61 mm aperture diameter, 80 mm focal depth focused transducer was simulated at the center of each face and oriented perpendicular to the scalp along the face’ normal. Acoustic simulations were performed at 200 kHz, 500 kHz and 1000 kHz using the well validated open-source hybrid angular spectrum solver mSOUND, which models acoustic propagation in high-contrast, heterogenous scattering media in seconds [22,23]. Those frequencies were chosen as they span the range of operating frequencies used in neurostimulation studies. The inputs of those simulations were acoustic maps derived from porosity maps *p* computed from CT (reference), our DL approach and pseudo-CT methods as implemented by Burgos et al. [35] and Izquierdo et al. [34] as follows:

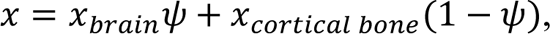

where *x* is a generic acoustic parameter standing for the speed-of-sound (*c*), attenuation (α) or density (ρ). The extreme values of those parameters corresponding brain and cortical bone tissues were extracted from the IT’IS Foundation acoustic parameter database [54]: *c*_*brain*_ = 1546 m/s and *c*_*cortical bone*_ = 3514 m/s, α_*brain*_ = 0.36 dB/(MHz · cm) and α_*cortical bone*_ = 7.74 dB/(MHz · cm), ρ_*brain*_ = 1046 kg/*m*^3^ and ρ_*cortical bone*_ = 1908 kg/*m*^3^. mSOUND propagates the acoustic energy along the z-axis of the input volume, therefore porosity maps were interpolated in the frame of the transducer using the rigid transformation aligning the transducer normal onto the transducer’ axis. mSOUND simulations were performed on a regular grid with resolution λ⁄5, where λ is the wavelength in water, thus resulting in longer simulations at high frequencies. Acoustic intensity maps were then interpolated back into the subject’ frame for all transducer positions. Finally, scalp maps were created by assigning to each face of the scalp mesh the total acoustic intensity deposited in the target brain region under study, in this work the left amygdala and the left thalamus.

## 3. Results

### 3.1 DL modifications that improved porosity estimates

Fig. 3 & 4 summarize the results of our systematic network optimization approach: Fig. 3 shows the DL features that had a significant impact on the accuracy of porosity estimates and Fig. 4 shows those that did not. Specifically, Fig. 3A shows that 2D ResNet with a 256×256 MRI T1w input significantly outperformed 2D UNet, which was true whether or not pCT was used as an additional input channel. We conclude that ResNet outperforms UNet for our estimation task and that pCT does not add significantly more information to the T1w input. Of all the network input sizes considered in this work, Fig. 3B shows that the highest performing configuration is 3D ResNet with 64×64×64 input patches (this result is consistent across all the performance metrics: MAE, MSE, PSNR and SSIM, see Table S1). This indicates that there may be more contextual information in small 3D patches than in larger 2D slices for the estimation of porosity. Fig. 3C shows that data augmentation significantly improved the estimation of porosity maps, although it is interesting to note that it is the inclusion of rotated version of the inputs that helped, not mirror flips. Finally, Fig. 3D shows that backpropagation in the mask (BIM) yielded a moderate but significant performance improvement, a result that is consistent across most other metrics (Table S2).

**Fig. 3.**
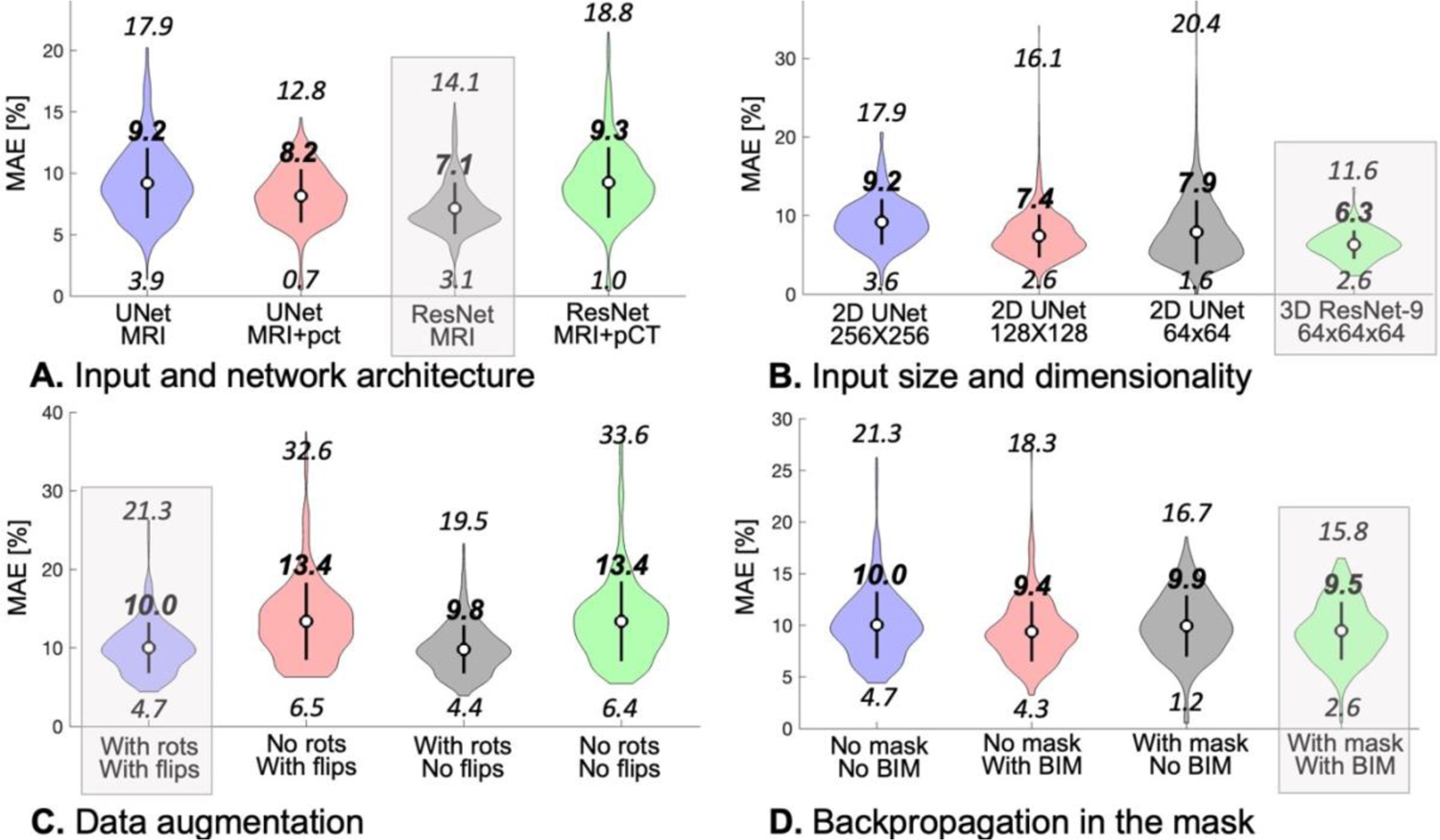
Violin plots of the mean absolute error (MAE) in the test set for various DL variations that were found to significantly impact the accuracy of porosity estimates. The bold numbers are the mean MAE for each variation, while the italic numbers above and below indicate the 99^th^ and 1^st^ percentiles of the error distribution, respectively. The grey boxes indicate the implementation variant retained in the final model. **A:** Impact of network architecture and input type. MRI + pCT indicates that both the MRI and the pseudo-CT (Izquierdo et al.) are used as input to the models (two channels). All inputs are 2D with sizes 256×256. **B:** Impact of input size and dimensionality. The 3D ResNet with 64×64×64 inputs has the best performance, therefore we retain it in the final model. **C:** Augmenting the data with rotations systematically improved the robustness of the estimation, whereas image flips had minimal impact. Nevertheless, we chose to perform both rotations and flips during training of the final model (comparisons performed for the 2D 256×256 UNet). **D:** Limiting the backpropagation from pixels in the skull mask improved the error distribution, therefore we retained this strategy in the final model (comparisons performed for the 2D 256×256 Unet).

**Fig. 4.**
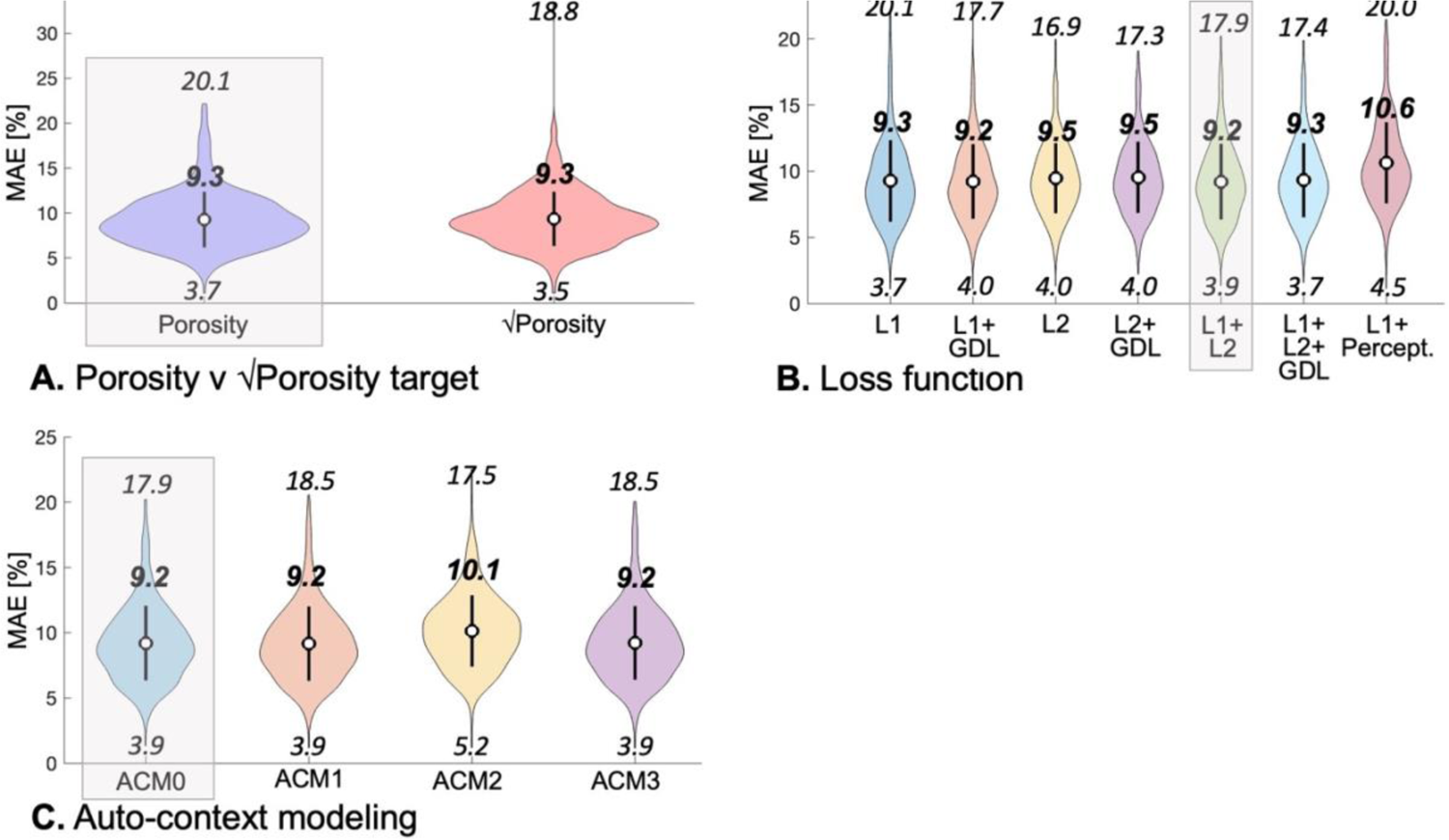
Violin plots of the mean absolute error (MAE) in the test set for various DL variations that did not significantly impact the accuracy of porosity estimates. The bold numbers indicate the mean MAE for each variation, while the italic numbers above and below indicate the 99^th^ and 1^st^ percentiles of the error distribution, respectively. The grey boxes indicate the implementation variant retained in the final model. **A:** Impact of porosity v square-root of the porosity as target estimation metric. Using the square-root of the porosity theoretically boosts small porosity values in the training process but in practice does not affect the training error much. **B:** All loss functions evaluated in this work yielded similar network estimation error across the test set, therefore we used a simple L1+L2 norm in the final implementation. **C:** Auto-context modeling did not improve estimation accuracy for our task, therefore we did not retain this technique in the final model (ACM0).

### 3.2 DL modifications that did not improve porosity estimates

Fig. 4A shows that using the square root of the porosity as the estimation target, which we thought could help boost small porosity values that may otherwise be under-represented in the MAE metric, did not significantly affect performance (this result is robust across all metrics, see Table S3). In Fig. 4B, the choice of the loss function beyond the common L1 and L2 norms was also found to have little impact. Specifically, the use of GDL and perceptual losses was found to only have a minor impact on final estimates, a result that is consistent across all metrics (Table S4). Finally, auto-context modeling was not found to improve estimates, therefore we did not retain it in the final implementation (Fig. 4C).

### 3.3 Training set size

Fig. 5 shows that the final training error for the 2D UNet 256×256 plateaued when using more than 9 subjects in the training set. This indicates that the size of the training set used in this work (13 subjects) is well matched to the complexity of the DL model.

**Fig. 5.**
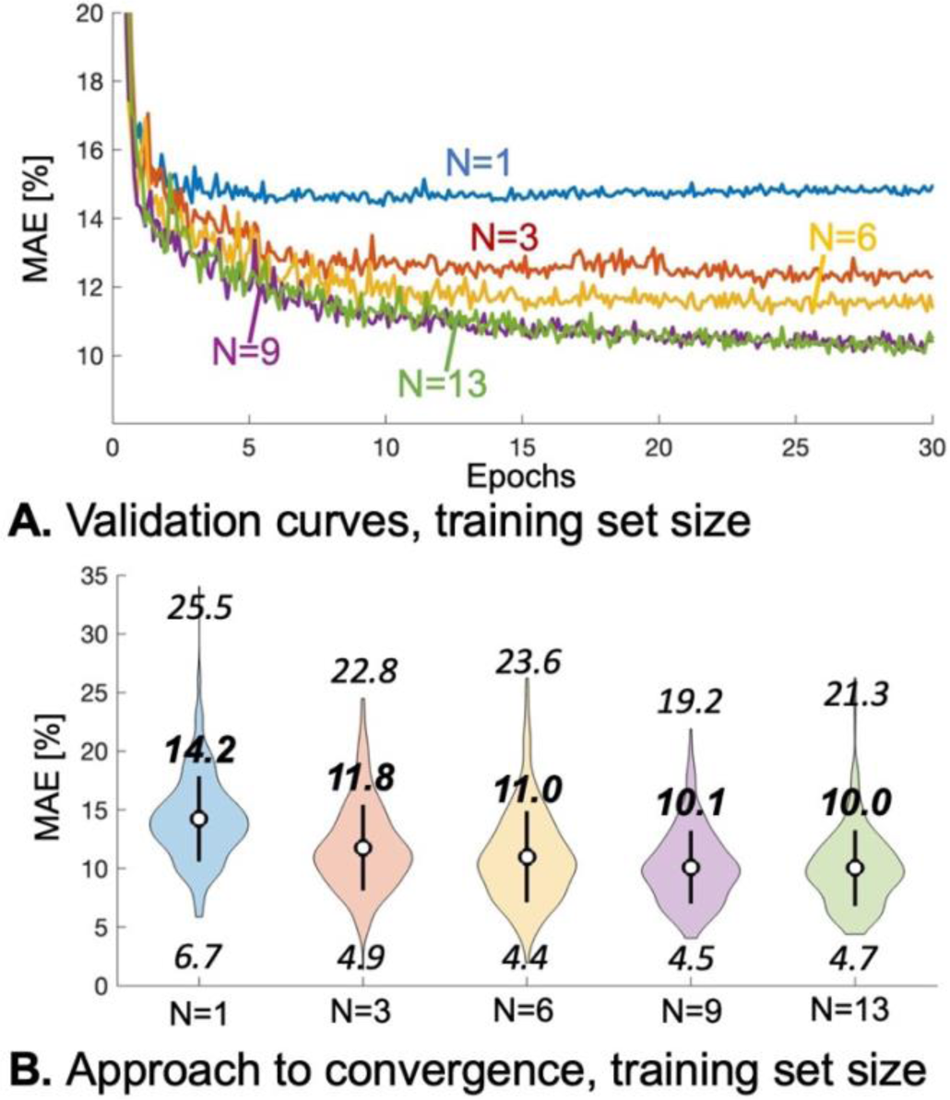
Impact of training set size on model performance. **A:** Validation curves for the baseline model (2D UNet, 256×256 inputs) trained with an increasing number of subjects. There are N=15 subjects in totals, one is set aside for validation while one is set aside for testing. **B:** Violin plot of the MAE error in the training set, as a function of the number of subjects in the training set. There is no significant performance improvement for N greater than 9, suggesting that using N=13 is well matched to the complexity of the model.

### 3.4 Proposed model

Based on the tests summarized in Figs. 3 & 4, we propose the following model: 3D ResNet cGAN with 64×64×64 input patches, L1+ L2 loss, backpropagation in the mask and data augmentation during training. Fig. 6 shows MAE error distributions in the test dataset for pix2pix, an out-of-the-box 2D 256×256 UNet cGAN [43], the pCT approach of Izquierdo et al. and our proposed DL. pCT and pix2pix have similar average error ∼9.4%, which is reduced to 6.9% using our optimized DL approach (a 26% reduction). Figs. 7 and S2 shows that although the average error of pCT is 9.4%, the error can be as high as 17% in some regions of the input volume. Using pix2pix resolves some of those problematic areas, as this strategy uses MRI-specific information whereas pCT only uses a population-average CT template. Our optimized DL approach yields even better overall estimation with few problem areas, although with a slight tendency to overestimate the porosity.

**Fig. 6.**
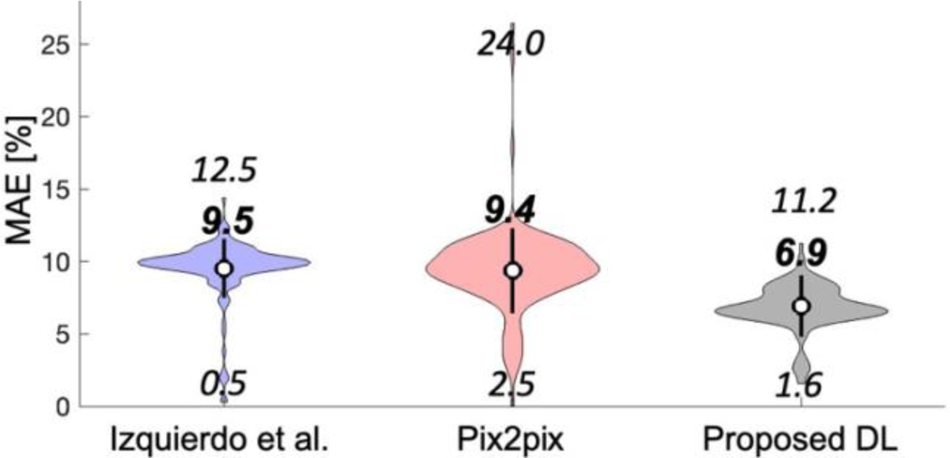
Final model performance, compared to pCT-based porosity estimation of Izquierdo et al. and the generic pix2pix GAN.

**Fig. 7.**
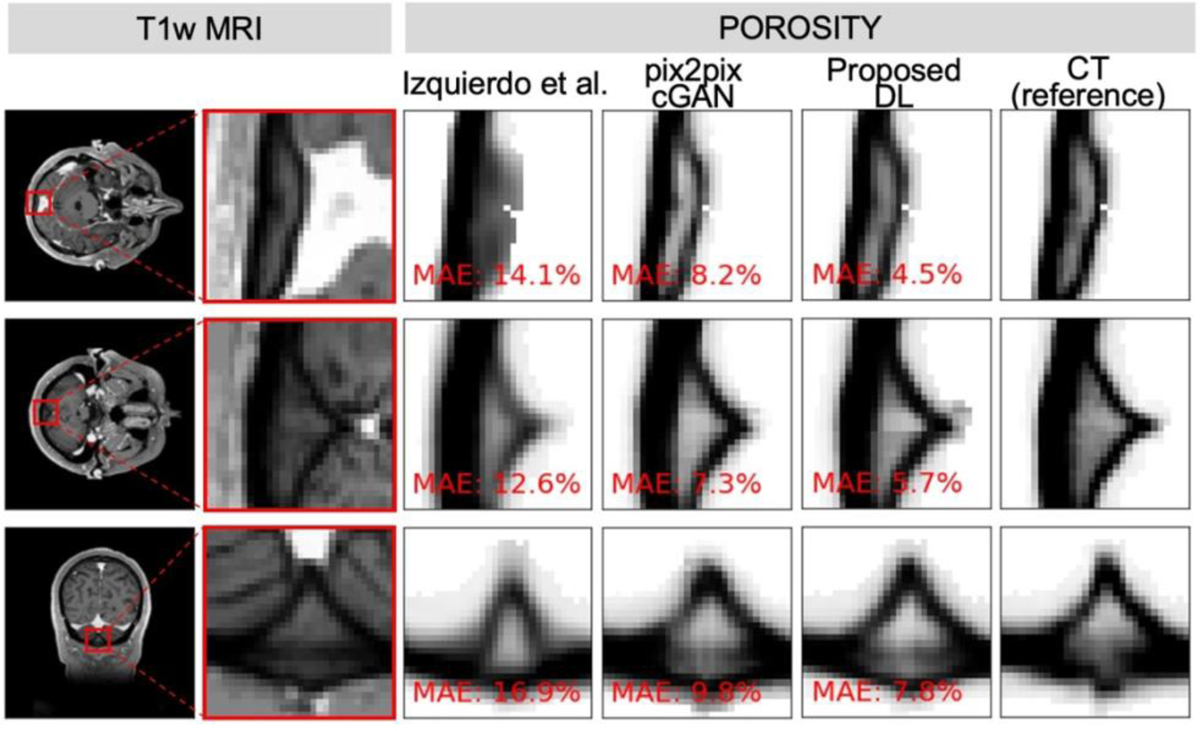
Representative examples of porosity maps estimated with the pCT (Izquierdo et al.), pix2pix, proposed DL approaches and compared to CT (reference). Also shown on the left are whole-FOV and zoom MRI slices, for reference.

### 3.5 Robustness to noise and sequence parameters

Fig. 8 shows porosity estimates in the test subject as a function of input image noise. Those results show that the proposed approach is robust to even very high noise levels in the T1-weighted image inputs. Indeed, the “very high noise” level corresponds to an SNR of 4 in the grey matter, which is extremely high compared to typical clinical image quality and is modeled here as an extreme test of the method.

**Fig. 8.**
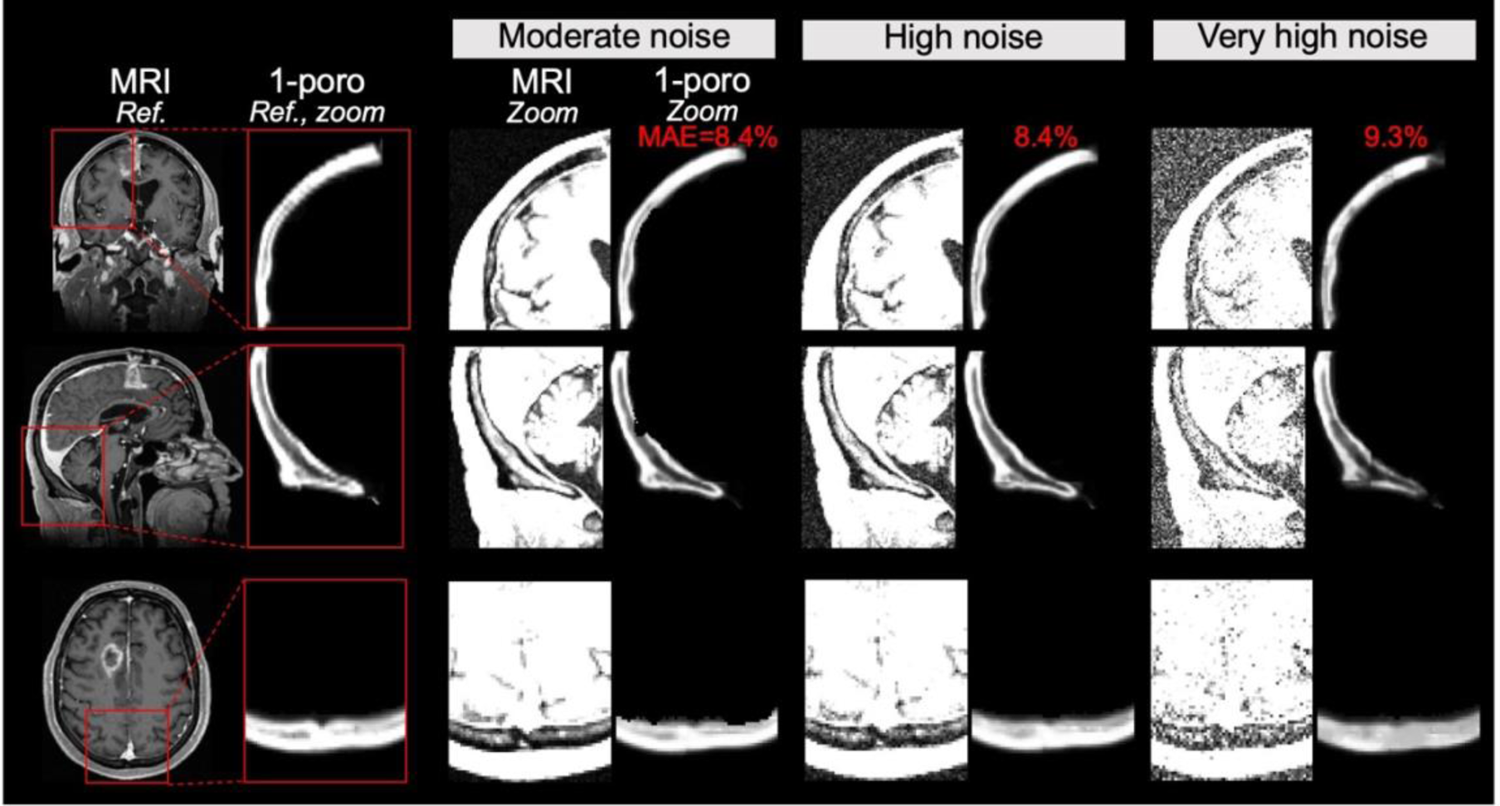
Performance of the proposed DL approach as a function of noise level in the input MRI image. The moderate, high and very high noise scenarios correspond to grey matter SNR values of 31, 10 and 4, respectively. For each noise level, we show zoomed panels of the noisy MRI inputs and the porosity image outputs. The mean absolute error (MAE) of porosity estimates is equal to 8.4%, 8.4% and 9.3% in the three noise scenarios, indicating that our proposed DL approach is robust to image noise.

Fig. 9 also shows that porosity estimation worked well even for subject data from the HCP and CERMEP database which were not present in the training/validation/test databases and were acquired with different acquisition parameters. This includes different sequence parameters, different coils and even different field strength for CERMEP. Estimations are reasonable in all subjects, specifically one can observe the expected high porosity values in large marrow-filled pores visible on the T1-weighted MRI inputs (Fig. 9, green arrows). Importantly, there were no “catastrophic failures”, where the estimation completely failed, for any of the HCP and CERMEP subjects, which can occur with other tools such as the pseudo-CT estimation approach of Burgos et al. The robustness of our approach to MRI acquisition parameters is likely due to the fact that estimation is restricted to inside the skull mask, which can be estimated robustly using SAMSEG/Freesurfer.

**Fig. 9.**
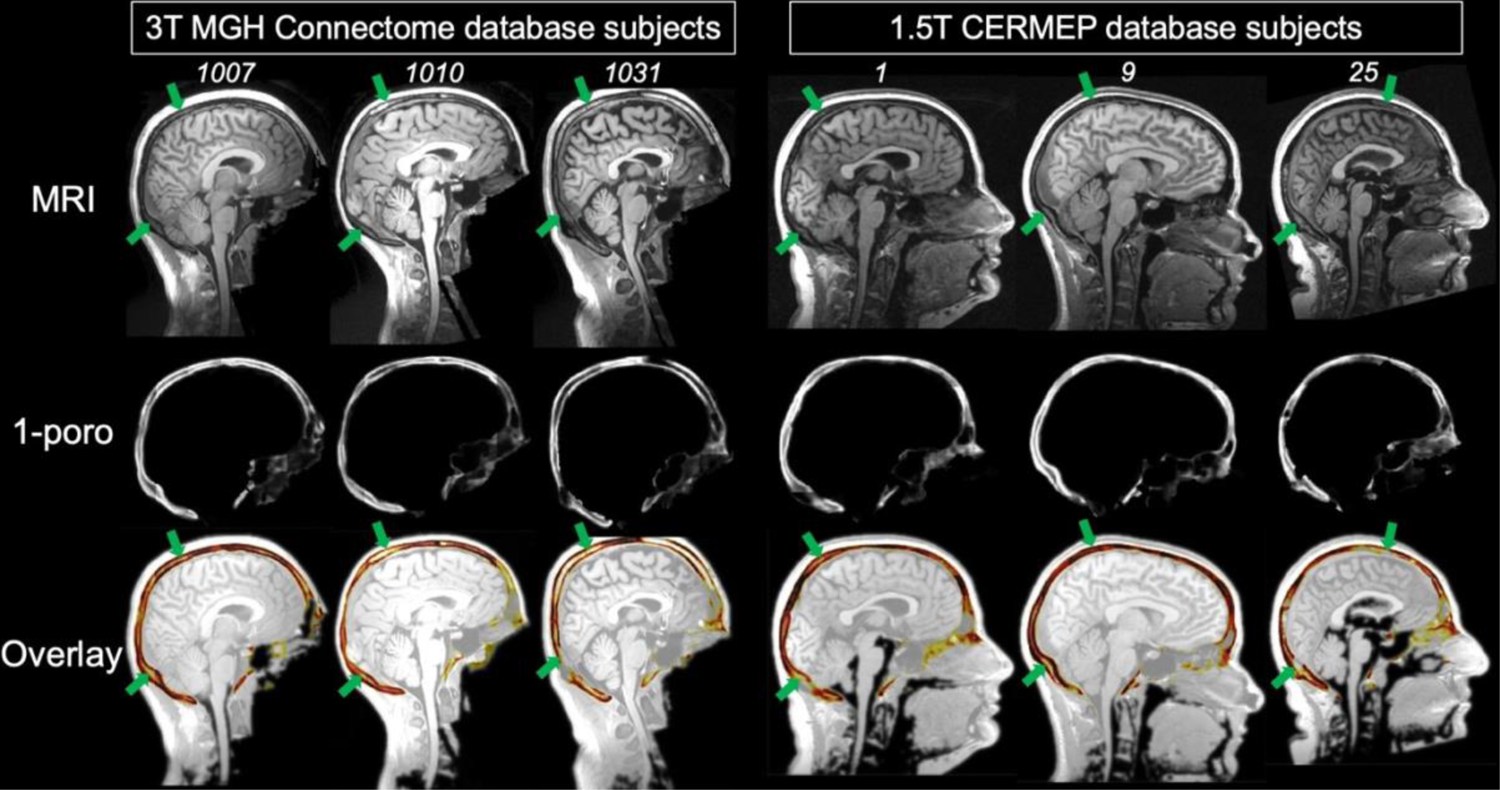
Porosity estimation using the proposed DL applied to subjects in the open-source Connectome and CERMEP databases, which were not included in the training, validation and testing. T1-weighted images from those databases were acquired on different MRI systems and with different sequence parameters than the inputs used for training, validation and testing. Estimates are reasonable for all subjects, with high porosity values corresponding to marrow-filled pores that are clearly visible on both the MRI and porosity images (green arrows).

### 3.6 Impact on acoustic simulations

Simulation of the 4868 transducer locations in the test subject using mSOUND took 8 min, 32 min and 121 min at 200 kHz, 500 kHz and 1000 kHz, respectively, with a parallelization factor of 20. Increasing computation time at increasing frequency is due to the increasing spatial resolution requirement to properly sample the decreasing wavelength (see Methods). Left thalamus scalp maps computed using CT-, DL-based and pCT-based porosity estimates, shown in Fig. 10, have similar relative distributions but different absolute amplitudes. The pCT approach of Izquierdo et al. led to optimal transducer position estimates in close agreement with those obtained using the reference CT data as well as our proposed DL approach. However, our DL approach yielded more accurate absolute dose estimates than when using the pCT method of both Burgos et al. and Izquierdo et al.

**Fig. 10.**
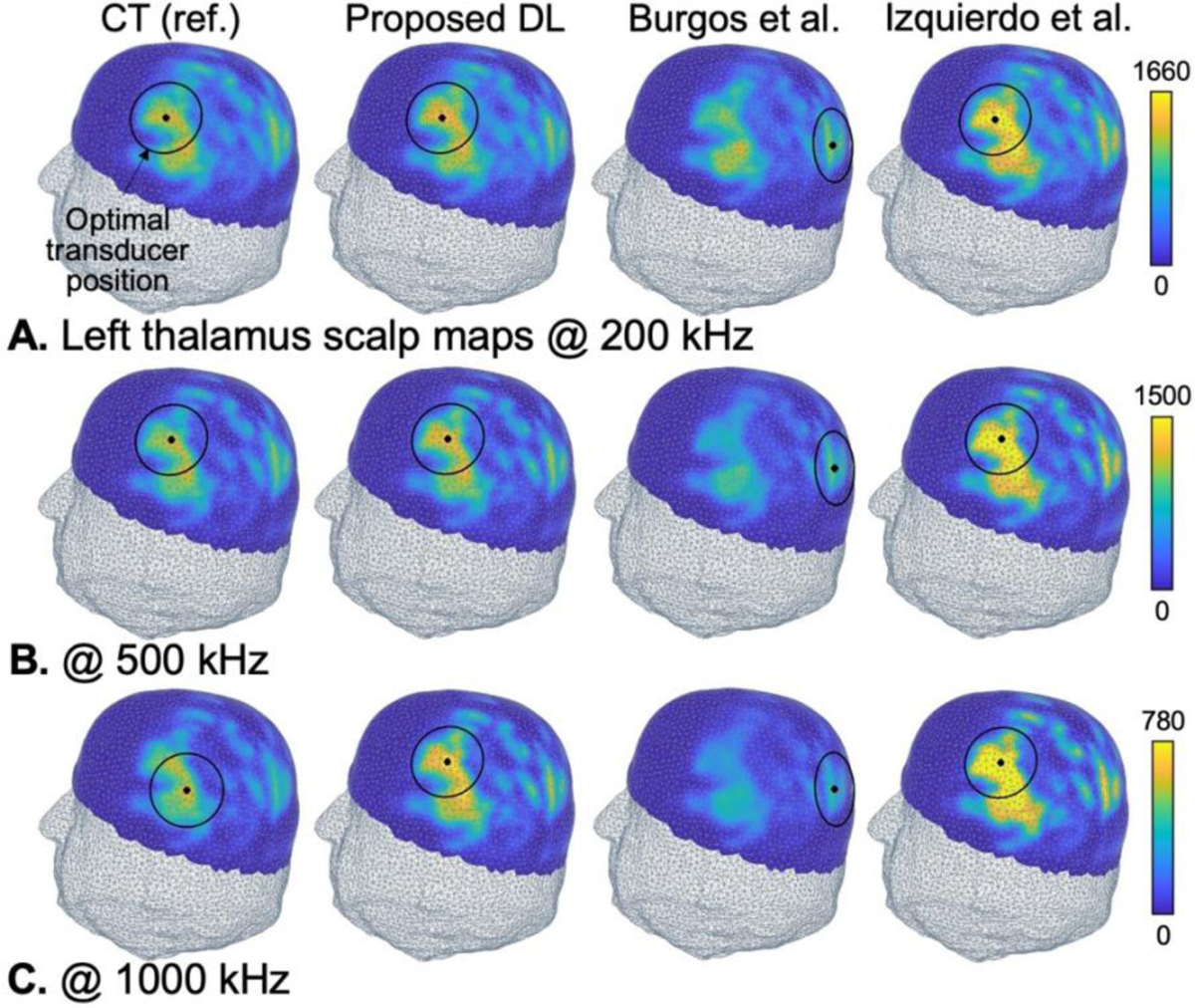
Scalp maps of the acoustic intensity deposited in the left thalamus of the test subject (arbitrary units), computed using mSOUND at 200 kHz, 500 kHz and 1000 kHz using porosity maps derived from CT (reference), the proposed DL approach and the pseudo-CT methods of Burgos et al. and Izquierdo et al.

## 4. Discussion

We optimized a deep learning (DL) strategy for estimation of skull porosity from MRI T1-weighted images, and deployed this approach for subject-specific neuronavigation of transcranial focused ultrasound (tFUS). Our approach is based on the observation that marrow-filled pores in the skull are visible on conventional T1- and T2-weighted MRI clinical images (Fig. 1). Although conventional MRI sequences do not provide skull contrast due to the short T2 of bone, most surrounding tissues are visible thus effectively yielding an inverted skull mask with the correct shape and thickness. Acoustic scattering in the head is largely determined by the skull shape, thickness and porosity; which led us to hypothesize that conventional T1w MR images contain all the information necessary for subject-specific acoustic simulations and that this information could be extracted using DL. An important aspect of our work is that we optimized the DL model by systematically assessing various implementation aspects for our specific estimation task. In contrast, most previous work implicitly assumed adequacy and optimality of architectures and implementation details. Among the DL strategies implemented and tested here, we found that the most impactful were the use of ResNet-9 compared to UNet (23% improvement in the MAE metric), 3D 64×64×64 input patches over 2D 256×256 slices (32% MAE improvement) and data augmentation with rotations and image flips (27% MAE improvement). Another strategy that yielded significant but smaller improvement was the use of backpropagation in the mask (BIM, 6% MAE improvement), a technique proposed in this work that consists in only including voxels falling in the skull mask during backpropagation. We hypothesized that this would improve accuracy and convergence as this focuses the network degrees-of-freedom on the voxels of importance as voxels outside the mask have a trivial porosity value equal to one. When testing BIM, we assessed several implementation approaches with and without masking the input and/or output and found that the best approach consists in not masking the input, but masking the output. This is likely because masking the input maximizes the amount of information passed to the network, while masking the output emphasizes learning on skull voxels only.

The best performing network (i.e., proposed approach) is a cGAN with a ResNet-9 generator and a patchGAN discriminator, 3D input patches of size 64×64×64 (1 mm isotropic), L1+L2 losses, backpropagation in the mask and data augmentation with rotations and image flips. This model improved estimation of the porosity map by 27% (MAE metric) compared to both pseudo-CT and pix2pix, a general pixel-to-pixel translation cGAN that we used as the baseline for our model [43]. However, those average errors do not tell the full story. As can be observed in Fig. 6, the tail of the error distribution associated with our DL approach is significantly shorter than tails associated with the pCT approach of Izquierdo et al. and pix2pix, thus indicating that outliers with estimation errors greater than 10% are largely resolved with our approach. This can be observed in Figs. 7 and S2, which show that errors associated with pCT-Izquierdo in small regions of the skull can be as high as ∼17%, which is largely resolved using our DL approach (∼6% error for the same regions). This represents a 25% improvement over pix2pix and a 65% improvement compared to pCT. We point out that atlas-based pseudo-CT approaches such as those by Izquierdo et al. and Burgos et al. were not designed for individualized porosity estimations and instead reflect, by definition, average distributions conserved across subjects. Those methods were developed for attenuation correction of positron emission tomography data, a task for which they were shown to be adequate as individualized porosity estimation is not important in this case [34]. This work shows that atlas-based pseudo-CT approaches are less appropriate for acoustic simulation since such simulations require accurate subject-specific porosity estimates that can differ significantly from the population average.

The DL strategies that we found did not significantly affect the results included using pCT as an additional input channel, using the square root of the porosity as the learning target (instead of the porosity); the specific loss function used during training and auto-context modeling (ACM). The fact that enriching the input with pCT or previous instances of the model (ACM) did not improve learning is somewhat at odd with previous work [46], and may therefore be specific to our estimation task. Finally, although our dataset was limited to 15 subject pairs of MRI and CT-based porosity 3D volumes, our results indicate that adding subjects beyond the first 9 did not significantly improve estimation results (Fig. 5). This is likely due to the fact that we divide each subject’ volumes in dozens of small 3D input patches, which decreases network complexity while increasing the size of the training set, and that we limit training in the skull mask (BIM), which focuses learning in the region of maximum importance.

An important result of this work is that the proposed DL approach was found to be robust to high levels of noise in the input images as well as varying MRI acquisition parameters, including sequence parameters, coils and even field strength. We attribute some of this robustness to our limitation of the estimation process to voxels inside the skull mask (BIM). Since estimation of the skull mask using SAMSEG/Freesurfer is itself a robust process, BIM acts as a sort of regularization by focusing the network solely on voxels directly associated with the estimation tasks (i.e. those in the skull). This removes the influence of voxels in the brain and the cerebrospinal fluid that, in the best of cases, likely have very little impact of the final estimate and, in the worst cases, would channel noise and unexpected contrast variation in the network output. In other words, BIM reduces overfitting and makes the estimation more robust by forcing the network to focus on voxels directly related to the estimation task. As a result, we believe that our trained network reason can be broadly useful to investigators in the field and therefore share it freely for download on GitHub (https://github.com/bastpg).

Relative distributions of the porosity were well estimated by our tool, however absolute porosity values were slightly overestimated compared to the reference values derived from CT. This led to a 5-22% overestimation of the dose in the brain in our acoustic simulations (error increases with frequency). It would be simple enough to correct this systematic bias using a global correction factor, however such inaccuracy is likely benign compared to the much greater errors introduced by imperfect porosity-to-acoustic parameter scaling. Indeed, published CT-to-speed-of-sound scaling laws have a 37-72% dispersion in the Hounsfield value range corresponding to cortical bone, while CT-to-acoustic attenuation laws have a 164-200% dispersion in the relevant range [12]. The cause of the large variability in the published Hounsfeld-to-acoustic parameter scaling relationships is currently unknown, although it is clear that some of it is caused by normal variations across subjects. In any case, such large variability likely leads to significant errors in the estimation of acoustic parameter which in turn affect all acoustic simulations, irrespective of the specific modeling tool employed. One possible solution could be to combine our DL-based porosity estimation approach with a limited number of ultrasound transmission measurements, since such measurements contain subject-specific information about the speed-of-sound (wave packet delay) and acoustic attenuation (transmission signal amplitude). We are currently investigating this approach, which well beyond the scope of the present work however.

We evaluated the impact porosity estimation errors on tFUS neuronavigation and dose estimation by modeling thousands of transducer positions on the scalp of a test subject using acoustic parameter maps derived from CT, pCT and the proposed DL approach. We summarize those simulations using ‘scalp maps’ showing the total acoustic energy deposited in the target brain region of interest for thousands of test transducer positions on the scalp of the subject. Scalp maps derived with our DL approach were close to those derived from CT data (both the relative and absolute dose distributions), and errors were larger with the pCT methods of Burgos et al and Izquierdo et al, which, as explained in a previous paragraph, is to be expected as atlas-based pCT methods cannot reflect individual porosity variations differing significantly from the population average. Our DL approach and the pCT approach of Izquierdo et al. yielded optimal transducer placement estimates that were very close to the reference treatment plan computed using CT data, although our DL approach yielded more accurate estimates of the absolute dose for all transducer locations. The treatment plan computed using the pCT approach of Burgos et al. yielded both greater transducer placement and absolute dose estimation errors, which shows that the implementation details and reference atlases used in different pCT methods have significant impact on tFUS simulations.

A limitation of our approach is that we only use T1-weighted MR images as inputs, however T2-weighted images also have positive bone marrow contrast and using them as additional input channels could further improve porosity estimates. In particular, our approach had a tendency to overestimate the porosity (i.e. errors are almost always an overestimation), which may be mitigated by using an additional T2-weighted input channel.

## 5. Conclusion

In this study we optimized a deep learning (DL) approach for estimation of skull porosity from T1-weighted MR input images. We found that using a ResNet generator was beneficial (versus UNet) along with using 3D input patches of size 64×64×64 (versus 2D patches of size 256×256 and 128×128). We also found that backpropagation in the mask (BIM), a technique that we propose here and consists in only including skull pixels in the backpropagation process, also improved the estimation. In contrast, using an additional pseudo-CT input channel and daisy-chaining DL networks (auto-context modeling) did not improve the estimation. We also found that the specific choice of the loss function had a small impact on model performance, and therefore use a simple L1+L2 loss to train the final model. The proposed model estimates porosity maps with ∼7% error, which yields near-optimal tFUS navigation transducer placement compared to CT-based modeling (reference) in the 200-1000 kHz frequency range as assessed using scalp maps of the acoustic dose delivered in the left amygdala and thalamus. The proposed approach is robust to input image noise and yields reasonable results for T1-weighted images acquired with a range of sequence parameters, coils and even field strengths, which makes it broadly useful. We share the trained weights on GitHub for maximum impact and dissemination.

## Data Availability

All data produced in the present study are available upon reasonable request to the authors

## Acknowledgements

This work was supported by a SPARC award from the McCance Center for Brain Health at the Massachusetts General Hospital, as well as NIH grants R01MH128421 and P41EB030006. The authors thank Yun Jing for helpful conversations about mSOUND.

## References

[1] W. Paulus, Transcranial brain stimulation: potential and limitations, ENeuroforum 5 (2014) 29–36. 10.1007/s13295-014-0056-6.

[2] A. Bystritsky, A.S. Korb, P.K. Douglas, M.S. Cohen, W.P. Melega, A.P. Mulgaonkar, A. Desalles, B.K. Min, S.S. Yoo, A review of low-intensity focused ultrasound pulsation, Brain Stimul 4 (2011) 125–136. 10.1016/j.brs.2011.03.007.

[3] C. Sarica, J.F. Nankoo, A. Fomenko, T.C. Grippe, K. Yamamoto, N. Samuel, V. Milano, A. Vetkas, G. Darmani, M.N. Cizmeci, A.M. Lozano, R. Chen, Human Studies of Transcranial Ultrasound neuromodulation: A systematic review of effectiveness and safety, Brain Stimul 15 (2022) 737–746. 10.1016/j.brs.2022.05.002.

[4] S.S. Yoo, A. Bystritsky, J.H. Lee, Y. Zhang, K. Fischer, B.K. Min, N.J. McDannold, A. Pascual-Leone, F.A. Jolesz, Focused ultrasound modulates region-specific brain activity, Neuroimage 56 (2011) 1267–1275. 10.1016/j.neuroimage.2011.02.058.

[5] Y. Tufail, A. Matyushov, N. Baldwin, M.L. Tauchmann, J. Georges, A. Yoshihiro, S.I.H. Tillery, W.J. Tyler, Transcranial Pulsed Ultrasound Stimulates Intact Brain Circuits, Neuron 66 (2010) 681–694. 10.1016/j.neuron.2010.05.008.

[6] P.P. Ye, J.R. Brown, K.B. Pauly, Frequency dependence of ultrasound neurostimulation in the mouse brain, Ultrasound Med Biol 42 (2016) 1512–1530.

[7] R.L. King, J.R. Brown, W.T. Newsome, K.B. Pauly, Effective parameters for ultrasound-induced in vivo neurostimulation, Ultrasound Med Biol 39 (2013) 312–331.

[8] H.A.S. Kamimura, S. Wang, H. Chen, Q. Wang, C. Aurup, C. Acosta, A.A.O. Carneiro, E.E. Konofagou, Focused ultrasound neuromodulation of cortical and subcortical brain structures using 1.9 MHz, Med Phys 43 (2016) 5730–5735.

[9] R.L. King, J.R. Brown, K.B. Pauly, Localization of ultrasound-induced in vivo neurostimulation in the mouse model, Ultrasound Med Biol 40 (2014) 1512–1522.

[10] K. Hynynen, J. Sun, Trans-skull ultrasound therapy: The feasibility of using image-derived skull thickness information to correct the phase distortion, IEEE Trans Ultrason Ferroelectr Freq Control 46 (1999) 752–755. 10.1109/58.764862.

[11] S.A. Leung, T.D. Webb, R.R. Bitton, P. Ghanouni, K. Butts Pauly, A rapid beam simulation framework for transcranial focused ultrasound, Sci Rep 9 (2019) 1–11. 10.1038/s41598-019-43775-6.

[12] S.A. Leung, D. Moore, T.D. Webb, J. Snell, P. Ghanouni, K. Butts Pauly, Transcranial focused ultrasound phase correction using the hybrid angular spectrum method, Sci Rep 11 (2021) 6532.

[13] K. Yoon, W. Lee, P. Croce, A. Cammalleri, S.S. Yoo, Multi-resolution simulation of focused ultrasound propagation through ovine skull from a single-element transducer, Phys Med Biol 63 (2018). 10.1088/1361-6560/aabe37.

[14] T. Deffieux, E.E. Konofagou, Numerical study of a simple transcranial focused ultrasound system applied to blood-brain barrier opening, IEEE Trans Ultrason Ferroelectr Freq Control 57 (2010) 2637–2653.

[15] J.K. Mueller, L. Ai, P. Bansal, W. Legon, Numerical evaluation of the skull for human neuromodulation with transcranial focused ultrasound, J Neural Eng 14 (2017) 066012.

[16] C. Baron, J.-F. Aubry, M. Tanter, S. Meairs, M. Fink, Simulation of intracranial acoustic fields in clinical trials of sonothrombolysis, Ultrasound Med Biol 35 (2009) 1148–1158.

[17] U. Vyas, D. Christensen, Ultrasound beam propagation using the hybrid angular spectrum method, in: 2008 30th Annual International Conference of the IEEE Engineering in Medicine and Biology Society, IEEE, 2008: pp. 2526–2529.

[18] S.L. Johnson, D.A. Christensen, C.R. Dillon, A. Payne, Validation of hybrid angular spectrum acoustic and thermal modelling in phantoms, International Journal of Hyperthermia 35 (2018) 578–590.

[19] U. Vyas, D. Christensen, Ultrasound beam simulations in inhomogeneous tissue geometries using the hybrid angular spectrum method, IEEE Trans Ultrason Ferroelectr Freq Control 59 (2012) 1093–1100. 10.1109/TUFFC.2012.2300.

[20] S. Almquist, D. Parker, D. Christensen, Simulation of hemispherical transducers for transcranial HIFU treatments using the hybrid angular spectrum approach, J Ther Ultrasound 3 (2015) 1–2.

[21] M. Hansen, D. Christensen, A. Payne, Experimental validation of acoustic and thermal modeling in heterogeneous phantoms using the hybrid angular spectrum method, International Journal of Hyperthermia 38 (2021) 1617–1626.

[22] J. Gu, Y. Jing, A modified mixed domain method for modeling acoustic wave propagation in strongly heterogeneous media, J Acoust Soc Am 147 (2020) 4055–4068. 10.1121/10.0001454.

[23] Y. Jing, M. Tao, J. Cannata, An improved wave-vector frequency-domain method for nonlinear wave modeling, IEEE Trans Ultrason Ferroelectr Freq Control 61 (2014) 515–524.

[24] K. Wang, E. Teoh, J. Jaros, B.E. Treeby, Modelling nonlinear ultrasound propagation in absorbing media using the k-Wave toolbox: experimental validation, in: 2012 IEEE International Ultrasonics Symposium, IEEE, 2012: pp. 523–526.

[25] B.E. Treeby, B.T. Cox, k-Wave: MATLAB toolbox for the simulation and reconstruction of photoacoustic wave fields, J Biomed Opt 15 (2010) 21314.

[26] E. Martin, J. Jaros, B.E. Treeby, Experimental validation of k-wave: Nonlinear wave propagation in layered, absorbing fluid media, IEEE Trans Ultrason Ferroelectr Freq Control 67 (2019) 81–91.

[27] F. Marquet, M. Pernot, J.F. Aubry, G. Montaldo, L. Marsac, M. Tanter, M. Fink, Non-invasive transcranial ultrasound therapy based on a 3D CT scan: Protocol validation and in vitro results, Phys Med Biol 54 (2009) 2597–2613. 10.1088/0031-9155/54/9/001.

[28] G. Bouchoux, K.B. Bader, J.J. Korfhagen, J.L. Raymond, R. Shivashankar, T.A. Abruzzo, C.K. Holland, Experimental validation of a finite-difference model for the prediction of transcranial ultrasound fields based on CT images, Phys Med Biol 57 (2012) 8005.

[29] S.A. Leung, D. Moore, Y. Gilbo, J. Snell, T.D. Webb, C.H. Meyer, G.W. Miller, P. Ghanouni, K. Butts Pauly, Comparison between MR and CT imaging used to correct for skull-induced phase aberrations during transcranial focused ultrasound, Sci Rep 12 (2022). 10.1038/s41598-022-17319-4.

[30] G.W. Miller, M. Eames, J. Snell, J. Aubry, Ultrashort echo-time MRI versus CT for skull aberration correction in MR-guided transcranial focused ultrasound: In vitro comparison on human calvaria, Med Phys 42 (2015) 2223–2233.

[31] E.M. Johnson, U. Vyas, P. Ghanouni, K.B. Pauly, J.M. Pauly, Improved cortical bone specificity in UTE MR Imaging, Magn Reson Med 77 (2017) 684–695.

[32] S. Guo, J. Zhuo, G. Li, D. Gandhi, M. Dayan, P. Fishman, H. Eisenberg, E.R. Melhem, R.P. Gullapalli, Feasibility of ultrashort echo time images using full-wave acoustic and thermal modeling for transcranial MRI-guided focused ultrasound (tcMRgFUS) planning, Phys Med Biol 64 (2019). 10.1088/1361-6560/ab12f7.

[33] F. Wiesinger, M. Bylund, J. Yang, S. Kaushik, D. Shanbhag, S. Ahn, J.H. Jonsson, J.A. Lundman, T. Hope, T. Nyholm, Zero TE-based pseudo-CT image conversion in the head and its application in PET/MR attenuation correction and MR-guided radiation therapy planning, Magn Reson Med 80 (2018) 1440–1451.

[34] D. Izquierdo-Garcia, A.E. Hansen, S. Förster, D. Benoit, S. Schachoff, S. Fürst, K.T. Chen, D.B. Chonde, C. Catana, An SPM8-based approach for attenuation correction combining segmentation and nonrigid template formation: application to simultaneous PET/MR brain imaging, Journal of Nuclear Medicine 55 (2014) 1825–1830.

[35] N. Burgos, M.J. Cardoso, K. Thielemans, M. Modat, S. Pedemonte, J. Dickson, A. Barnes, R. Ahmed, C.J. Mahoney, J.M. Schott, Attenuation correction synthesis for hybrid PET-MR scanners: application to brain studies, IEEE Trans Med Imaging 33 (2014) 2332–2341.

[36] H. Liu, M.K. Sigona, T.J. Manuel, L.M. Chen, B.M. Dawant, C.F. Caskey, Evaluation of Synthetically Generated CT for use in Transcranial Focused Ultrasound Procedures, ArXiv Preprint ArXiv:2210.14775 (2022).

[37] F. Sammartino, D.W. Beam, J. Snell, V. Krishna, Kranion, an open-source environment for planning transcranial focused ultrasound surgery, J Neurosurg 132 (2019) 1249–1255.

[38] M. Miscouridou, J.A. Pineda-Pardo, C.J. Stagg, B.E. Treeby, A. Stanziola, Classical and learned MR to pseudo-CT mappings for accurate transcranial ultrasound simulation, IEEE Trans Ultrason Ferroelectr Freq Control 69 (2022) 2896–2905.

[39] P. Su, S. Guo, S. Roys, F. Maier, H. Bhat, E.R. Melhem, D. Gandhi, R. Gullapalli, J. Zhuo, Transcranial MR imaging–guided focused ultrasound interventions using deep learning synthesized CT, American Journal of Neuroradiology 41 (2020) 1841–1848.

[40] S.N. Yaakub, T.A. White, E. Kerfoot, L. Verhagen, A. Hammers, E.F. Fouragnan, Pseudo-CTs from T1-weighted MRI for planning of low-intensity transcranial focused ultrasound neuromodulation: An open-source tool, Brain Stimulation: Basic, Translational, and Clinical Research in Neuromodulation 16 (2023) 75–78.

[41] H. Koh, T.Y. Park, Y.A. Chung, J.H. Lee, H. Kim, Acoustic Simulation for Transcranial Focused Ultrasound Using GAN-Based Synthetic CT, IEEE J Biomed Health Inform 26 (2022) 161–171. 10.1109/JBHI.2021.3103387.

[42] O. Puonti, J.E. Iglesias, K. Van Leemput, Fast and sequence-adaptive whole-brain segmentation using parametric Bayesian modeling, Neuroimage 143 (2016) 235–249.

[43] P. Isola, J.-Y. Zhu, T. Zhou, A.A. Efros, Image-to-image translation with conditional adversarial networks, in: Proceedings of the IEEE Conference on Computer Vision and Pattern Recognition, 2017: pp. 1125–1134.

[44] A. Chapman, G. Ter Haar, Thermal ablation of uterine fibroids using MR-guided focused ultrasound-a truly non-invasive treatment modality, Eur Radiol 17 (2007) 2505–2511.

[45] O. Ronneberger, P. Fischer, T. Brox, U-net: Convolutional networks for biomedical image segmentation, in: Medical Image Computing and Computer-Assisted Intervention–MICCAI 2015: 18th International Conference, Munich, Germany, October 5-9, 2015, Proceedings, Part III 18, Springer, 2015: pp. 234–241.

[46] D. Nie, R. Trullo, J. Lian, L. Wang, C. Petitjean, S. Ruan, Q. Wang, D. Shen, Medical image synthesis with deep convolutional adversarial networks, IEEE Trans Biomed Eng 65 (2018) 2720–2730.

[47] C. Wang, C. Xu, C. Wang, D. Tao, Perceptual adversarial networks for image-to-image transformation, IEEE Transactions on Image Processing 27 (2018) 4066–4079.

[48] B. Fischl, FreeSurfer, Neuroimage 62 (2012) 774–781.

[49] M. Reuter, H.D. Rosas, B. Fischl, Highly accurate inverse consistent registration: a robust approach, Neuroimage 53 (2010) 1181–1196.

[50] J.-F. Aubry, M. Tanter, M. Pernot, J.-L. Thomas, M. Fink, Experimental demonstration of noninvasive transskull adaptive focusing based on prior computed tomography scans, J Acoust Soc Am 113 (2003) 84–93.

[51] K. Ugurbil, D. Van Essen, HCP Young Adult, https://www.Humanconnectome.org/Study/Hcp-Young-Adult (n.d.).

[52] I. Mérida, J. Jung, S. Bouvard, D. Le Bars, S. Lancelot, F. Lavenne, C. Bouillot, J. Redouté, A. Hammers, N. Costes, CERMEP-IDB-MRXFDG: a database of 37 normal adult human brain [18 F] FDG PET, T1 and FLAIR MRI, and CT images available for research, EJNMMI Res 11 (2021) 1–10.

[53] Q. Fang, D.A. Boas, Tetrahedral mesh generation from volumetric binary and grayscale images, Proceedings - 2009 IEEE International Symposium on Biomedical Imaging: From Nano to Macro, ISBI 2009 (2009) 1142–1145. 10.1109/ISBI.2009.5193259.

[54] IT’IS Foundation, IT’IS Foundation Material Property Database, https://Itis.Swiss/Virtual-Population/Tissue-Properties/Database/ (n.d.).

