## Supplementary material for "Robust deep learning estimation of cortical bone porosity from MR T1-weighted images for individualized transcranial focused ultrasound planning"

**Table S1**

DL performance for different input patch dimensions and generator architectures. All models are trained with L1+L2 loss, masked output porosity, backpropagation in the mask and 60 epochs.

| | **MAE** | **MSE** | **PSNR** | **SSIM** |
|---|---|---|---|---|
| **2D UNet** 256x256 | 9.3% ± 2.6% | 3.3% ± 1.6% | 34.2 ± 5.7 | 96.8 ± 1.2 |
| **2D UNet** 128x128 | 10.2% ± 4. 4% | 3.3% ± 2.8% | 34.2 ± 7.0 | 91.2 ± 5.5 |
| **2D UNet** 64x64 | 14.3% ± 8.3% | 6.0% ± 6.6% | 28.2 ± 9.3 | 85.9 ± 12.8 |
| **3D ResNet** 64x64x64 | **7.7% ± 3.8%** | **1.9% ± 2.4%** | **39.8 ± 7.4** | **92.5 ± 6.7** |

**Table S2**

DL performance with and without backpropagation in the mask (BIM). All models are 2D 256x256 cGANs with a UNet generator, 2D PatchGAN discriminator, L1 loss and trained over 30 epochs.

| | **MAE** | **MSE** | **PSNR** | **SSIM** |
|---|---|---|---|---|
| **No mask / No BIM** | 10.0% ± 2.9% | 3.9% ± 0.2% | 32.4 ± 5.3 | 95.6% ± 1.4% |
| **No mask / With BIM** | **9.4% ± 2.8%** | **3.4% ± 0.2%** | **33.9 ± 5.4** | 96.2% ± 1.5% |
| **With mask / No BIM** | 10.3% ± 2.8% | 4.0% ± 0.2% | 32.3 ± 5.2 | 95.1% ± 1.5% |
| **With mask / With BIM** | 9.8% ± 2.8% | 3.6% ± 0.2% | 33.1 ± 5.3 | **96.2% ± 1.3%** |

**Table S3**

DL performance for porosity and square root of the porosity target estimation outputs. Errors are computed with respect to the porosity map in each case. The model is a 2D 256x256 cGAN with a UNet generator, 2D PatchGAN discriminator, L1 loss, backpropagation in the mask (masked), 30 epochs trainings.

| | **MAE** | **MSE** | **PSNR** | **SSIM** |
|---|---|---|---|---|
| **Porosity** | **9.3% ± 2.8%** | 3.5% ± 0.2% | 33.7 ± 5.8 | 96.6 ± 1.3 |
| **Sqrt porosity** | 9.4% ± 2.6% | **3.4% ± 0.1%** | **33.9 ± 5.4** | **96.7 ± 1.1** |

**Table S4**

DL performance for different training losses. All models are 2D 256x256 cGANs with a UNet generator, 2D PatchGAN discriminator, L1 loss, backpropagation in the mask (masked), 30 epochs trainings.

| Model | MAE | MSE | PSNR | SSIM |
|---|---|---|---|---|
| **L1** ($\boldsymbol{\lambda_1 = 1, \lambda_{disc} = 0.01}$) | 9.3% ± 2.8% | 3.5% ± 0.2% | 33.7% ± 5.8% | **96.6% ± 1.3%** |
| **L1 + GDL** ($\boldsymbol{\lambda_1 = 0.5, \lambda_3 = 0.5,\ \lambda_{disc} = 0.01}$) | 9.3% ± 2.6% | 3.3% ± 0.1% | 34.2% ± 5.6% | 96.2% ± 1.4% |
| **L2** ($\boldsymbol{\lambda_2 = 1, \lambda_{disc} = 0.01}$) | 9.5% ± 2.5% | **3.2% ± 0.1%** | **34.5% ± 5.3%** | 96.5% ± 1.3% |
| **L2 + GDL** ($\boldsymbol{\lambda_2 = 0.5, \lambda_3 = 0.5,\ \lambda_{disc} = 0.01}$) | 9.6% ± 2.6% | 3.2% ± 0.1% | 34.4% ± 5.4% | 96.2% ± 1.4% |
| **L1 + L2** ($\boldsymbol{\lambda_1 = 0.5, \lambda_2 = 0.5, \lambda_{disc} = 0.01}$) | **9.3% ± 2.6%** | 3.3% ± 0.1% | 34.3% ± 5.7% | 96.5% ± 1.3% |
| **L1 + L2 + GDL** ($\boldsymbol{\lambda_1 = 0.33, \lambda_2 = 0.33,\ \lambda_3 = 0.33,\ \lambda_{disc} = 0.01}$) | 9.4% ± 2.6% | 3.2% ± 0.1% | 34.3% ± 5.6% | 96.4% ± 1.3% |
| **L1 + Perceptual** ($\boldsymbol{\lambda_1 = 0.5, \lambda_4 = 0.5, \lambda_{disc} = 0.01}$) | 10.8% ± 2.7% | 3.9% ± 0.2% | 32.4% ± 5.2% | 93.5% ± 2.1% |

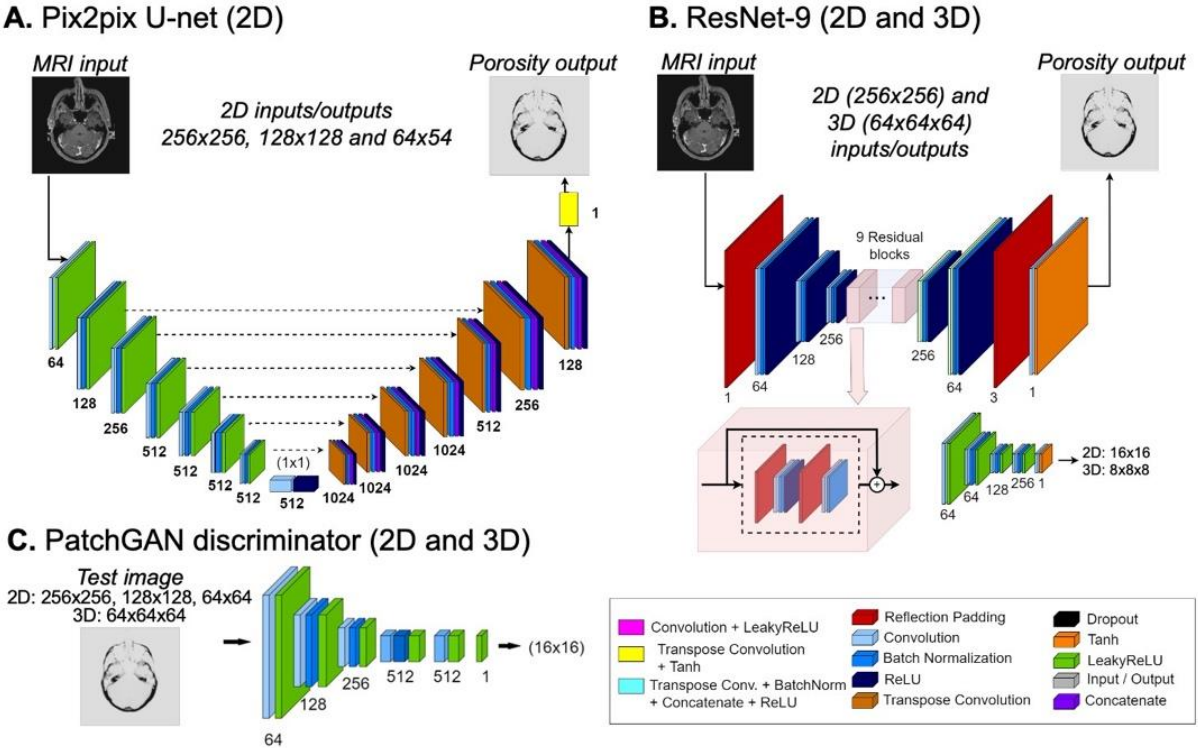


Fig. S1. Generator and discriminator networks. **A:** 2D ‘pix2pix’ UNet generator from Isola et al. (2016). The original network accommodates inputs/outputs of size 256x256, which we modified to 128x128 and 64x64 by removing pairs of downsampling/upsampling blocks. All networks employ an image of size 1x1x512 at the bottom of the ‘U’. **B:** ResNet with 9 residual block layers generator. 2D and 3D versions were implemented accommodating inputs/outputs of size 256x256 and 64x64, respectively. **C:** PatchGAN discriminator. The same network was used for all 2D input sizes. Therefore, discriminator output size varied with the input size, and was 16x16 for 256x256 inputs, 8x8 for 128x128 inputs and 4x4 for 64x64 inputs. A 3D version of PatchGAN was also implemented with a 4x4x4 outputs for 64x64x64 inputs.

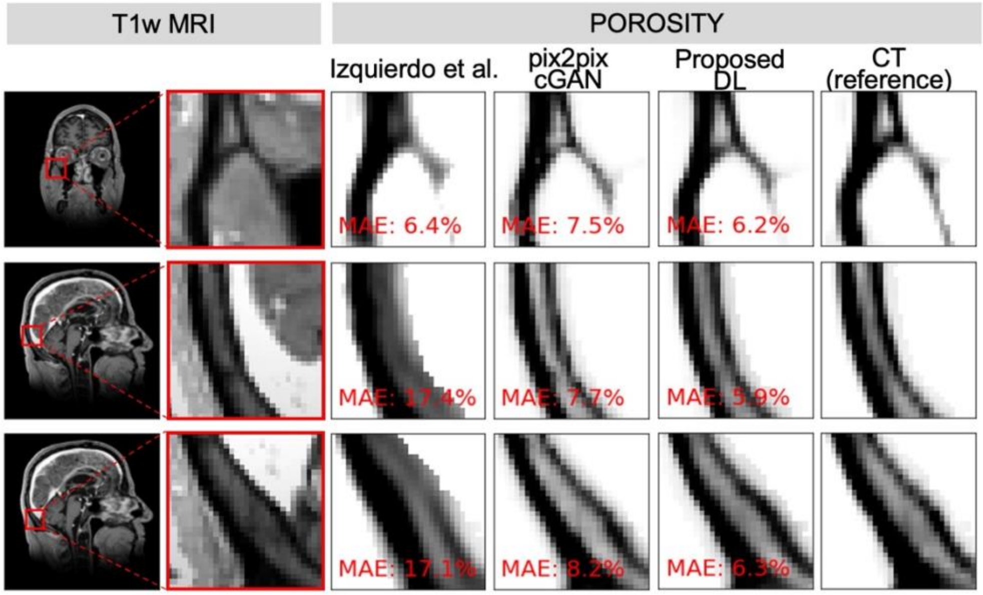


Fig. S2. Representative examples of porosity maps estimated with the pCT (Izquierdo et al.), pix2pix, proposed DL approaches and compared to CT (reference). Also shown on the left are whole-FOV and zoom MRI slices, for reference.

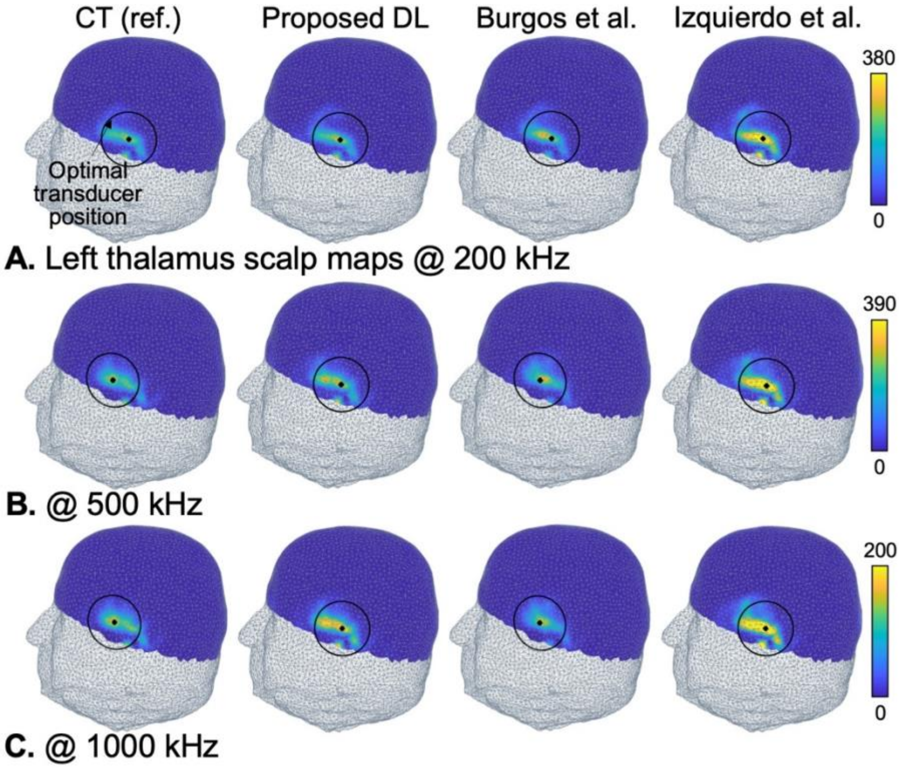


Fig. S3. Scalp maps of the acoustic intensity deposited in the left amygdala of the test subject (arbitrary units), computed using mSOUND at 200 kHz, 500 kHz and 1000 kHz using porosity maps derived from CT (reference), the proposed DL approach and the pseudo-CT methods of Burgos et al. and Izquierdo et al.

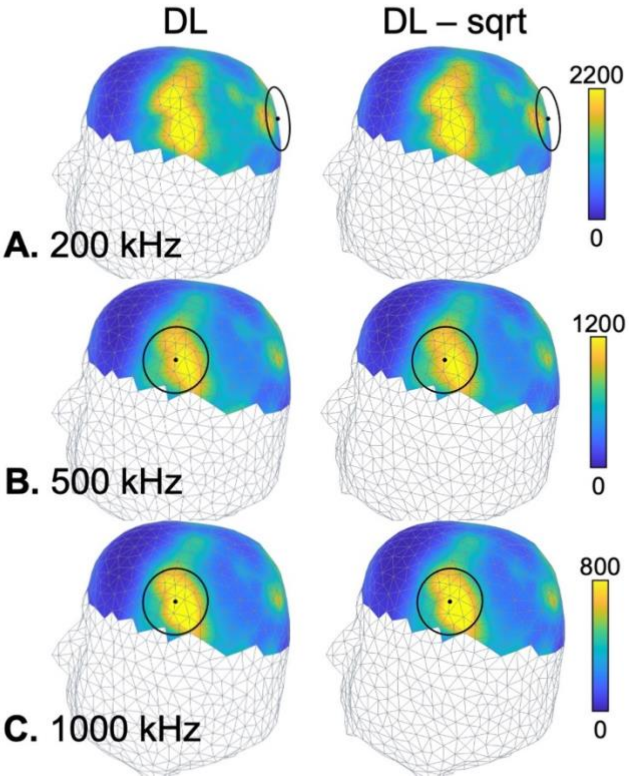


Fig. S4. Left thalamus scalp maps computed using mSOUND at 200 kHz, 500 kHz and 1000 kHz using porosity maps derived from DL networks trained on the porosity (left) and on the square root of the porosity (right). There are no noticeable differences between the two sets of maps.
